# Mindfulness-Oriented Recovery Enhancement rebalances prefrontal responses to drug and natural reward cues in opioid use disorder

**DOI:** 10.64898/2026.02.12.26346211

**Authors:** Yuefeng Huang, Ahmet O. Ceceli, Greg Kronberg, K Rachel. Drury, Sarah G. King, Natalie E. McClain, Yui Ying Wong, Maggie Boros, Eduardo R. Butelman, Pierre-Olivier Gaudreault, Muhammad A. Parvaz, Nelly Alia-Klein, Eric L. Garland, Rita Z. Goldstein

## Abstract

Despite decades of clinical implementation of medications for opioid use disorder (OUD), overdose mortality rates remain high, underscoring a critical gap in treatments that target brain mechanisms driving addiction. Mindfulness-Oriented Recovery Enhancement (MORE) has demonstrated efficacy in reducing opioid use and craving, hypothetically by restructuring the salience of drug and natural rewards. Yet, to date, MORE’s neurobiological mechanisms remain unclear. In this first functional magnetic resonance imaging (fMRI) randomized controlled trial (RCT) of MORE for OUD (NCT04112186), we tested whether compared with an active psychoeducational supportive therapy (PST) control group, MORE rebalanced neural responses to drug and natural reward cues in inpatients with OUD receiving standard of care including medications. Compared with PST, eight weeks of MORE significantly reduced drug-biased activity in the dorsolateral prefrontal cortex (dlPFC) and posterior regions of the default mode network including the precuneus during downregulation of responses to drug cues relative to upregulation of responses to natural reward cues (even when controlling for passive cue viewing). The shift from drug to natural reward responses in the lateral and ventromedial PFC was associated with lower cue-induced craving exclusively in the MORE group. MORE also reduced medial PFC synchronization to naturalistic drug-related movie scenes and significantly extended abstinence duration at follow-up (∼4 months post-treatment) relative to PST. Together, this neuroimaging RCT demonstrates that MORE normalizes function in PFC nodes of the reward, salience, and control systems, positioning MORE as a biologically-grounded adjunct to pharmacotherapy for OUD.

## Introduction

The opioid epidemic remains a major health crisis in the United States, with approximately 80,000 overdose deaths in 2023 alone involving opioids.^1^ Driven by heroin and fentanyl co-use, overdose is the leading cause of death for people between the ages 18 and 44.^2^ However, treatment options for opioid use disorder (OUD) are limited. Several medications for OUD, including buprenorphine and methadone, reduce illicit opioid use and lower mortality; yet their effectiveness is constrained by high attrition and relapse. Specifically, treatment dropout rates average around 30%, approximately 50% of patients treated with medications for OUD relapse within 6 months^3,4^, and 91% of individuals with OUD (iOUD) report a lapse within the first year post-treatment.^5–7^ Thus, many iOUD remain vulnerable for continued non-medical drug use, overdose, or death, underscoring the urgent need for adjunctive neuroscience-informed behavioral interventions that can be integrated with medications for OUD to improve treatment outcomes.

The impaired response inhibition and salience attribution (iRISA) model^8–11^ posits that individuals with substance use disorder, including OUD, exhibit heightened cortico-striatal reactivity to drug cues at the expense of non-drug cues including salient natural rewards, across brain networks involved in reward and salience processing such as the ventromedial prefrontal cortex (vmPFC), as well as in higher order executive control regions including the dorsolateral PFC (dlPFC). Indeed, our previous studies have shown that, as compared to healthy controls, iOUD exhibited hyperactivation of the vmPFC when viewing drug-related vs. neutral images.^12^ In these participants, we also reported increased orbitofrontal cortex synchronization (a measure of shared neural dynamics reflecting temporally aligned activity across individuals) when viewing a drug-themed movie (especially during drug-related scenes), which decreased after 15 weeks of inpatient treatment.^13^ Consistent with the iRISA model, this drug-biased PFC reactivity is evident even when drug cues are compared with non-drug natural rewards (e.g., food or sexual cues),^12,14^ suggesting dysregulated hedonic responses. Importantly, because increased drug craving and shorter abstinence duration in iOUD are associated with such higher drug cue-reactivity PFC signaling^12–15^, its reduction may serve as a key interventional target for OUD recovery.

Mindfulness-Oriented Recovery Enhancement (MORE) is an evidence-based behavioral intervention that uniquely integrates mindfulness with reappraisal and savoring practices to remediate the reward dysregulation underpinning addiction by shifting salience attribution away from drug cues and toward natural rewards.^16^ To date, MORE has demonstrated efficacy for reducing opioid use and misuse, as well as pain and craving, in seven randomized controlled trials (RCTs).^17^ For example, in a full-scale RCT of MORE for patients with opioid misuse (62% with a full OUD diagnosis) in primary care (N=250), MORE reduced opioid misuse by 45%, nearly tripling the effect of supportive psychotherapy (OR=2.94) at a 9-month follow-up.^18^ A second full-scale RCT in current and former military personnel with long term opioid use (N=230) replicated these effects, with MORE significantly reducing opioid use, craving, and illicit drug use relative to supportive psychotherapy.^19^ In a RCT of MORE as an adjunct to methadone treatment for people with OUD (N=154), adding MORE to standard addictions care resulted in 42% less drug relapse and 59% less treatment discontinuation, and fewer days of drug use, than usual addiction care without MORE.^20^ Studies investigating MORE’s psychophysiological underpinnings suggest that MORE treats addiction by reducing drug cue-reactivity and enhancing responsiveness to natural rewards. An electroencephalogram (EEG) study of MORE that estimated drug cue-reactivity via late positive potentials (LPP) during drug and neutral image presentation indicated higher drug cue-reactivity at baseline, which normalized following eight weeks of therapy only in the MORE group (whereas those in supportive psychotherapy showed sustained drug-related LPP).^21^ In this study, individuals in the MORE group also showed more effective downregulation of emotional reactivity to drug cues via reappraisal, and upregulation of emotional reactivity to natural reward cues via savoring; the latter mediated MORE’s relationship with reductions in opioid misuse at a 3-month follow-up. Similarly, a recent EEG study found that MORE normalized OUD-related hedonic dysregulation as indicated by increases in the LPP during savoring of natural rewards, which mediated the effect of MORE on reduced opioid craving.^22^

While these efforts illustrate the efficacy of MORE and highlight drug cue-reactivity and its regulation via reappraisal and savoring as potential treatment targets, the neural mechanisms underlying MORE’s therapeutic effects are less clear. The limited neurobiological evidence suggests that MORE is closely aligned with the iRISA model, targeting the cortico-striatal circuitry that underlies drug-biased cue-reactivity. In a sample of 13 healthy adult smokers with nicotine dependence, as compared to a control group that received no treatment (n=6), eight weeks of MORE significantly reduced smoking cue-reactivity and increased savoring of natural reward images via changes in ventral striatum and vmPFC activity; increases in savoring-related brain activity correlated with smoking reduction and increased positive affect.^16^ Similar patterns have been extended to women with OUD in another preliminary study (n=9), illustrating that eight weeks of MORE decreased emotion regulation difficulty and increased PFC-related functional connectivity during the MORE mindfulness practice.^23^ These neural changes point to potential mechanistic targets of MORE. In our previous larger-scale fMRI study, compared to healthy controls, treatment-seeking inpatient iOUD exhibited lateral PFC hyperactivations during reappraisal of drug images vs. savoring of food images, as associated with higher medication dosage (an index of addiction severity).^12^ Obtained earlier in treatment, these findings suggest that the effort needed to downregulate responses to drug cues via reappraisal may come at the expense of the ability to upregulate healthy hedonic responses, depleting the cognitive-affective resources needed to enjoy natural, non-drug rewards in addiction. Notably, anterior cingulate cortex activity during food savoring was correlated with longer treatment duration in iOUD, suggesting that successful treatment may restore natural reward processing.^12^ Plausibly, MORE may remediate these mechanisms to restore disrupted cortico-striatal reward, salience, and control processes in iOUD.

To date, no fMRI-based RCT has yet identified the neural substrates by which MORE (or other mindfulness-based interventions) influence drug cue-reactivity, reappraisal, and savoring in OUD (or any other substance use disorder). In this RCT, we aimed to examine the effects of eight weeks of MORE compared to eight weeks of psychoeducational supportive therapy (PST) in treatment-seeking inpatient (heroin-primary) iOUD receiving standard of care including medications for OUD. Neural outcomes focused on the regulation of cortico-striatal drug cue-reactivity assessed using two well-documented task paradigms from our prior work, one involving passive viewing and reappraisal of drug-related static pictures,^12^ and the other involving passive viewing of a naturalistic drug-theme movie^13^. We also measured savoring of natural reward cues (i.e., food) in the former paradigm. In addition, we examined clinical outcomes, including abstinence length and drug craving ratings. Guided by the iRISA model and previous studies,^9,12,13^ we hypothesized that, compared to PST: a) MORE would facilitate downregulation of drug-biased cortico-striatal activity in iOUD, reflected in decreased drug cue-reactivity to static images and/or decreased drug-biased synchronization during movie watching, and b) rebalance reappraisal and savoring-related neural activity such that more PFC resources would be allocated to food cues (savoring) relative to drug cues (reappraising). Additionally, we expected that these neural changes would be accompanied by (and related to) improved clinical outcomes, including reduced drug craving and longer abstinence.

## Methods

### Participants

Participants were recruited as part of a clinical trial (NCT04112186). A total of 92 iOUD completed the baseline MRI, MRI1, in this study (Figure 1). After exclusions, 86 of these participants were assigned to eight weeks of MORE (n = 41) or PST (n = 45) via stratified block randomization (strata: sex and severity of dependence, block size=2) and 62 of them (MORE: n=26; PST n=36) came back for the second MRI, MRI2, after treatment completion. Due to the nature of behavioral interventions, participants and therapists were not blinded to treatment assignment; however, investigators, data analysts, and outcome assessors were blinded to group allocation to minimize assessment and analytic bias. The drug cue-reactivity task analyses included data from both MRI sessions provided by 59 participants (MORE: n = 24; PST: n = 35). The movie-watching task analyses were conducted in a subset of 37 participants (MORE: n = 18; PST: n = 19). Of the 59 participants, data were analyzed from 38 subjects (of 40) who returned for the follow-up visit (MORE: n = 13; PST: n = 25); the interval between MRI2 and follow-up was 115.76 ± 22.57 days in MORE and 115.96 ± 76.20 days in PST (i.e., ∼4 months post-treatment).

**Figure 1:**
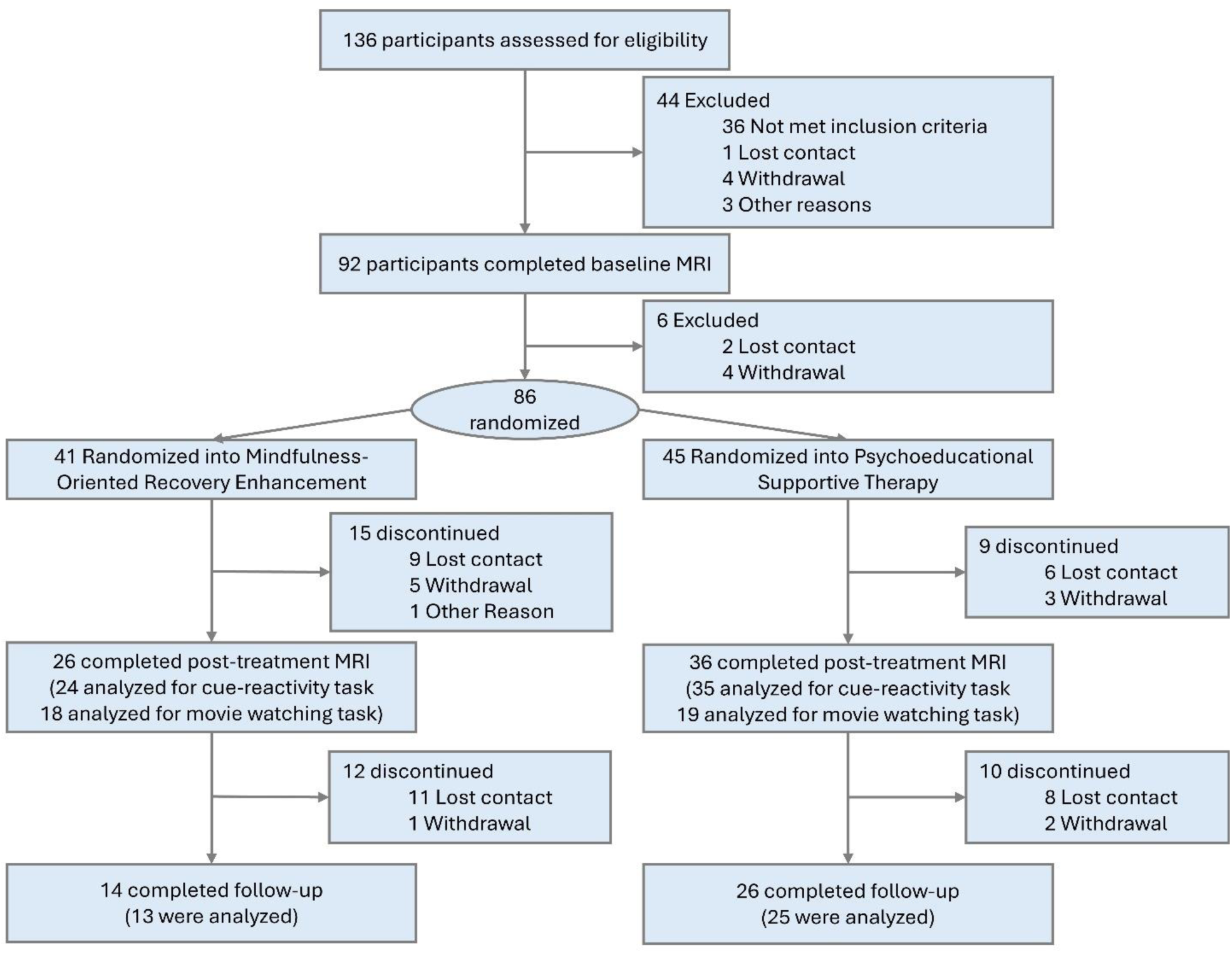
CONSORT Diagram Depicting the Study Flow. No significant differences in age, sex, education, race, ethnicity, verbal and nonverbal IQ, smoking status, age of onset for opioid use or treatment length at baseline emerged between the treatment groups. Depression symptoms and severity of drug dependence decreased after treatment in both groups. See table 1 for details.

**Table 1.** Sample Profile.

| Variable | MORE<br>(N=24) | PST<br>(N=35) | Significance test |
| --- | --- | --- | --- |
| Demographic |  |  |  |
| Age, years | 43.64 (11.29) | 42.75 (9.99) | t(57) = 0.32, p = .75 |
| Sex (men/women) | 19/5 | 27/8 | $\chi^2_1$ (N= 59) = 0.00, p = 1.00 |
| Education, years | 11.79 (1.74) | 12.34 (2.70) | W = 331, p = .15 |
| Race (white/black/other) | 16/1/7 | 21/7/7 | p = .18 |
| Ethnicity (Hispanic/non-Hispanic) | 9/15 | 13/22 | $\chi^2_1$ (N= 59) = 0.00, p = 1.00 |
| Verbal IQ | 95.75 (12.40) | 96.11 (12.37) | t(57) = 0.11, p = .91 |
| Nonverbal IQ | 9.67 (3.40) | 10.11 (2.76) | t(57) = 0.56, p = .58 |
| Baseline only drug-related variables |  |  |  |
| Smoker<br>(non/past/current) | 0/0/24 | 0/3/32 | $\chi^2_1$ (N= 59) = .76, p = .39 |
| Age of onset for opioid use | 22.33 (6.84) | 24.25 (8.22) | W = 309.5, p = .64 |
| Treatment length at baseline MRI scan day | 146.3 (123.5) | 194.8 (158.9) | t(56) = 1.23, p = .22 |
| Dynamic mood-related variables |  |  |  |
| BDI (pre-treatment) | 13.21 (10.75) | 12.66 (10.39) | Group main effect: F(1, 56.20) = 0.76, p = .39<br>Time main effect: F(2, 89.49) = 4.54, p = .013*<br>Interaction effect: F(2, 89.49) = 2.79, p = .067 |
| BDI (post-treatment) | 10.26 (10.38) | 9.52 (9.06) |  |
| BDI (follow-up) | 11.00 (9.95) | 7.17 (7.71) |  |
| BAI (pre-treatment) | 11.46 (8.39) | 9.23 (9.45) | Group main effect: F(1, 56.28) = 0.98, p = .33<br>Time main effect: F(2, 90.57) = 3.04, p = .053<br>Interaction effect: F(2, 90.57) = 0.26, p = .77 |
| BAI (post-treatment) | 10.29 (10.43) | 8.85 (8.93) |  |
| BAI (follow-up) | 7.15 (6.28) | 5.25 (8.46) |  |
| Dynamic drug-related variables |  |  |  |
| SDS (pre-treatment) | 10.78 (4.14) | 11.43 (3.67) | Group main effect: F(1, 54.15) = 0.67, p = .42<br>Time main effect: F(1, 50.13) = 10.92, p = .002*<br>Interaction effect: F(1, 50.13) = 0.02, p = .90 |
| SDS (follow-up) | 7.92 (4.87) | 8.72 (4.56) |  |
| SOWS (pre-treatment) | 5.95 (7.73) | 2.11 (3.03) | Group main effect: F(1, 91) = 5.12, p = .026<br>Time main effect: F(1, 91) = 0.86, p = .36<br>Interaction effect: F(1, 91) = 1.81, p = .18 |
| SOWS (follow-up) | 3.54 (3.04) | 2.56 (4.49) |  |
| HCQ (pre-treatment) | 40.23 (11.43) | 38.00 (16.07) | Group main effect: F(1, 50.14) = 3.63, p = .063<br>Time main effect: F(1, 37.50) = 4.00, p = .053<br>Interaction effect: F(1, 37.50) = 2.59, p = .12 |
| HCQ (follow-up) | 39.00 (9.76) | 28.12 (8.11) |  |
| SMAST (pre-treatment) | 4.24 (4.21) | 2.14 (3.00) | Group main effect: F(1, 39.88) = 2.32, p = .14<br>Time main effect: F(1, 29.73) = 3.42, p = .074<br>Interaction effect: F(1, 29.73) = 2.70, p = .11 |
| SMAST (follow-up) | 2.23 (1.42) | 1.74 (2.56) |  |
| FTND (pre-treatment) | 3.45 (1.47) | 3.32 (2.10) | Group main effect: F(1, 58.10) = 0.02, p = .90<br>Time main effect: F(1, 48.00) = 0.01, p = .92<br>Interaction effect: F(1, 48.00) = 0.16, p = .69 |
| FTND (follow-up) | 3.38 (2.50) | 3.46 (2.17) |  |
| Heroin use in past 30 days (pre-treatment) | 0.21 (0.41) | 0.17 (0.71) | Group main effect: $F(1, 60.50) = 0.96, p = .33$ |
| Heroin use in past 30 days (post-treatment) | 0.96 (3.32) | 0.63 (2.41) | Time main effect: $F(2, 101.68) = 1.66, p = .19$ |
| Heroin use in past 30 days (follow-up) | 1.15 (4.16) | 0.36 (1.08) | Interaction effect: $F(2, 101.68) = 0.46, p = .63$ |

\* Indicates significant results. Significant tests were examined with chi-squared tests, Fisher's exact tests, 2-sample t tests with equal or unequal variance, and Wilcoxon rank-sum tests, mixed model ANOVA with linear mixed models as appropriate. P values were Bonferroni-corrected for multiple comparisons within each variable set: demographic variables ( $0.05/7 = 0.007$ ), baseline drug-related variables ( $0.05/3 = 0.017$ ), dynamic mood-related variables ( $0.05/2 = 0.025$ ), and dynamic drug-related variables ( $0.05/6 = 0.008$ ). Values are presented as mean (SD) or N. follow-up = approximately 4-months after MRI2 MORE: Mindfulness-Oriented Recovery Enhancement; PST: Psychoeducational supportive therapy; Verbal IQ was measured with Wide Range Intelligence Test–III, standard score; Nonverbal IQ was measured with Wechsler Abbreviated Scale of Intelligence, Matrix Reasoning Scaled Score. BAI, Beck Anxiety Inventory; BDI, Beck Depression Inventory-II; FTND, Fagerström Test for Nicotine Dependence; HCQ, Heroin Craving Questionnaire; SDS, Severity of Dependence Scale; SMAST, Short Michigan Alcohol Screening Test; SOWS, Short Opiate Withdrawal Scale.

Participants were recruited from a medication-assisted inpatient treatment facility in the greater New York City area. Each underwent a comprehensive clinical diagnostic interview conducted by trained research staff under the supervision of a licensed clinical psychologist. All participants were medically stabilized on methadone or buprenorphine for at least two weeks prior to study participation and met Diagnostic and Statistical Manual of Mental Disorder (DSM)-5 criteria for OUD, with heroin as the primary drug of choice or reason for treatment. The clinical diagnostic interview encompassed the Mini-International Neuropsychiatric Interview^24^ for DSM-5 criteria and the Addiction Severity Index,^25^ a semi-structured instrument that captures drug use history and severity. Severity of drug dependence, craving, and withdrawal symptoms were assessed with the Severity of Dependence Scale,^26^ Heroin Craving Questionnaire (a modified version of the Cocaine Craving Questionnaire), ^27,28^ and Short Opiate Withdrawal Scale^29^ for all participants. A brief physical examination, including heart rate, blood pressure, urine drug toxicology, alcohol concentration (via saliva testing strips), and breath carbon monoxide levels, and a review of medical history were also performed by trained research staff. All women were tested for pregnancy via urine testing strips at each study visit/session day. See Supplementary Materials for a detailed breakdown of drug use and comorbidities for our participants. All participants provided written informed consent, and study procedures were approved by the institutional review board of the Icahn School of Medicine at Mount Sinai. Adverse events and unintended effects were monitored throughout the study; no serious adverse events related to the intervention were observed.

### Interventions

The empirically supported and manualized MORE intervention^17–20,30^ was delivered over eight weekly two-hour sessions. Sessions provided training in mindfulness meditation to strengthen meta-awareness and attentional control, reappraisal to decrease negative emotions and craving, and savoring to enhance natural reward responsiveness and positive emotion regulation. Led by at least master’s level therapists, participants were guided to use mindfulness practices to disrupt drug-related attentional bias and to savor the pleasant sensory features and positive feelings elicited by naturally rewarding objects and events. In addition to the weekly group sessions, participants were instructed to engage in daily 15-minute mindfulness, reappraisal, and savoring practice sessions using instructional recordings. The manualized active control condition in this study, PST, consisted of eight weekly two-hour group sessions involving psychoeducation and group support around addiction recovery-related topics pertinent to coping with stress, managing cravings, and addressing relapse triggers and adverse consequences of opioid use. PST participants were instructed to engage in 15-minutes of journaling a day on opioid-related themes. In order to prevent treatment diffusion, different therapists completed the MORE and PST arms in each cycle, and treatment fidelity was monitored throughout the trial. Therapists received formal training and supervision from the developer of MORE. On average, participants in the MORE group attended 5.7 ± 2.4 treatment sessions and the PST group attended 5.4 ± 2.5 sessions with no significant group differences [t(57) = 0.47, p = 0.64].

### MRI data acquisition

The MRI protocol was optimized to be Human Connectome Project compatible^31^ and the data was collected on a Siemens 3-T Skyra scanner (Siemens Healthcare, Erlangen, Germany) using a 32-channel head coil. See Supplementary Materials for details.

### Picture cue-reactivity task

#### Task paradigm and cue-induced craving ratings

The task paradigm was reported in detail in our previous studies.^12,32^ The task comprised of three conditions: participants were instructed to passively look at drug (images of drug preparation, use, and paraphernalia), food, and neutral images (i.e., *look* condition), actively downregulate their emotional reactivity to the drug images (i.e., *reappraise* condition), and upregulate their emotional reactivity to food images (i.e., *savor* condition). The task was administered over three 6 min 54 s functional runs.

As our a priori selected cue-induced drug craving measure, we chose to use the post-MRI craving ratings provided on half of the drug images viewed during the task; this choice was guided by our previous research^12,32^ where this specific craving measure correlated with dlPFC drug reappraisal activity. A 2 (treatment group: MORE vs. PST) by 2 (session: pre-vs. post-treatment) mixed ANOVA was conducted for the cue-induced craving ratings; see Supplementary Materials for detailed task instructions and these (and additional) task-related ratings.

#### Data preprocessing

Raw BOLD-fMRI data in DICOM (Digital Imaging and Communications in Medicine) format were first converted to NIFTI (Neuroimaging Informatics Technology Initiative) via dcm2nii^33^ and HeuDiConv (https://github.com/nipy/heudiconv) and adapted to the BIDS (Brain Imaging Data Structure) format^34^ and then preprocessed via fMRIprep pipeline (version 20.2.1).^35,36^ See Supplementary Materials for details.

#### BOLD fMRI data analyses

Individual parameter estimates were generated using the GLM via FMRIB Software Library (FSL)’s FEAT (version 6.0).^37^ The picture cue-reactivity task is a block design task and the GLM regressors included three “look” events, one “reappraise” event, and one “savor” event, sampled from the onset of the corresponding trials (16-second length, convolved with a double gamma hemodynamic response function). Translation and rotation parameters for motion (x, y, and z for each), global components of white matter and CSF, and motion outlier time points were included as “nuisance” regressors. Fixation events were not modeled, contributing to the implicit baseline. A fixed-effects model was used for subject-level statistical maps of each task event (look drug/food/neutral, reappraise drug, savor food) and their contrasts to yield estimates of drug cue-reactivity (1. look drug > look neutral or 2. look drug > look food), drug reappraisal (3. reappraise drug > look drug), food savoring (4. savor food > look food), and rebalancing of drug and food responses [5. reappraise drug > savor food and 6. (reappraise drug > look drug) > (savor food > look food) to account for valence and arousal of food and drug cues]. Group-level estimates were calculated using FSL FLAME 1+2 mixed-effects model to improve group-level variance estimation and population inferences via Markov chain Monte Carlo simulations.^38^ Guided by our previous research^12,32^ and the primary aims of the present study, we preselected the above-mentioned six contrasts of interest *a priori* to test our hypotheses. These contrast estimates were selected to evaluate the treatment group (MORE vs. PST) by session (pre-vs. post-treatment scan) interaction effects following FSL’s Common GLM design guidance (https://web.mit.edu/fsl_v5.0.10/fsl/doc/wiki/GLM.html). To inspect correlations between treatment-related changes in brain activity and drug craving, we first computed within-subject subtraction (post-minus pre-treatment, or Δ) of subject-level statistical mapping of the emotion regulation contrasts that showed significant interaction effects, as well as of our selected measure of cue-induced drug craving. Finally, we also examined whether the treatment-related changes in brain activity (and craving) were associated with abstinence duration at follow-up. All BOLD data analyses were conducted voxel-wise using a cluster-defining threshold of Z > 3.1^39^ and corrected to a cluster-extent threshold of p<0.05.

### Movie task

#### Task paradigm and scene-induced craving ratings

The task paradigm was reported in detail in our previous study.^13^ Participants passively viewed the first 17 min 3 sec of the movie “Trainspotting.” The video was played on a television outside the MRI bore, which participants viewed via a mirror mounted on the head coil, while MRI-compatible in-ear headphones were used for the audio. Thirty-four three-second movie clips were used to acquire scene-induced drug craving ratings within 45 minutes of movie viewing. This was our a priori craving measure of interest for the movie-related analyses.^13^ A 2 (treatment group: MORE vs. PST) by 2 (session: pre-vs. post-treatment scan) mixed ANOVA was conducted for the scene-induced craving ratings; see the Supplementary Materials for details and additional movie ratings.

#### Data preprocessing

Anatomical and functional data were preprocessed using fMRIPrep version 20.2.1, followed by custom preprocessing, global component regression, and an region of interest (ROI)-based shared response model applied per group^13^; see the Supplementary Materials for details.

#### Identifying synchronized repetition times and region-specific reactivity

Before and after treatment, within each ROI and group we aimed to identify individual repetition times exhibiting significant synchronization across participants, indicated by peaks in the group median of the shared component^13^. At each repetition time we estimated 95% confidence intervals for the group median via 5000 subject-wise bootstraps. We then compared this confidence interval for the median to the maximum value that could be expected by chance synchronization. We performed 5000 phase randomizations of the shared component [by taking the Fast Fourier Transform (FFT), randomizing the phase of all frequency components, and then taking the inverse FFT], calculated the median signal and kept the maximum value over all repetition times from each randomization to build a null distribution. A repetition time had a significant response if the 95% confidence interval for the group median was above the 95th percentile of the null distribution. This procedure yielded a set of significant repetition times, T*={t_i_}, which reflects synchronized activity within a given group of subjects in a specific ROI.

To gain insight into the movie content that drove the synchronization in a group of subjects, T*, we performed a reverse correlation-like analysis (inspired by analysis of single unit recordings) similar to Hasson et al.^40^ For each t_i_ in T*, we collected a 5 second clip of the movie corresponding to t_i_-10 to t_i_-5 to account for the delay in the hemodynamic response. Each 5 second clip was split into 1 second bins that were labeled drug/non-drug (see movie labeling). This procedure mapped T* to the fraction of unique time bins that were labeled as drug (out of total bins), which we called the drug bias and used as a test statistic. For contrasts between groups, p-values were obtained by taking the difference between the drug bias for each group and comparing this difference to a null distribution generated by randomizing T* for each group 5000 times. For treatment group (MORE vs. PST) by session (pre-vs. post-treatment scan) interaction effects, the test statistic was the difference in drug bias between sessions, compared between groups. To adjust for the number of ROIs in whole brain analyses, we applied FDR correction to the p-values for all ROIs. We also performed additional analyses with correction only to the number of ROIs within the PFC, guided by our previous studies.^12,13,32^

In summary, we quantified the drug cue-reactivity of each ROI by first identifying TRs during the movie with a significant response in that ROI across the group of subjects, reflecting synchronized brain responses to the movie. Drug cue-reactivity was then computed as the fraction of such TRs that followed movie drug content compared to non-drug content, accounting for the hemodynamic delay.^13^

#### Movie labeling

To map synchronized TRs to movie content, we first binned the movie into 1 second clips with the start and end of each clip aligned to a repetition time^13^. Each 1 second clip was then labeled by raters blind to the current analyses/results as drug if it contained any drugs, drug use, or a character who was intoxicated; and labeled as non-drug otherwise. This yielded 464 seconds labeled drug out of 1023 seconds total (45.4%).

#### ISC correlations

We used inter-subject correlations (ISC) as a subject-level measure to test for relationships between brain responses and behavioral outcomes (craving and abstinence)^13^. In this analysis, ISCs are derived by Pearson correlations between each subject’s ROI time course with the mean time course from the rest of the group (leave-one-subject-out). The group mean can be conceptualized as a model of iOUD brain dynamics in response to naturalistic drug cues in a given ROI. The ISC scores then reflect individual subjects’ expression of these canonical dynamics^41^ which we hypothesized to be related to our main clinical outcome variables (craving and abstinence). ISC scores were derived specifically in ROIs that showed a significant treatment group (MORE vs. PST) by session (pre-vs. post-treatment scan) interaction effect of the drug-biased synchronization. The ISC scores were then Fisher z-transformed, subtracted between sessions for each subject, and then inverse z-transformed to get a ΔISC score per subject. These ΔISC scores were then correlated with the Δscene-induced craving scores from the post-movie survey (averaged over all scenes) and abstinence at follow-up (corrected for abstinence at MRI2 and days between the MRI2 and follow-up visits) using Spearmen correlation.

### Abstinence duration analyses

Self-reported abstinence duration was collected at the pre- and post-treatment MRI days and at the follow-up session. These reports were manually checked and cross-referenced with self-reported days of drug use in the past 30 days and with urine toxicology results obtained on the day of each session.

To test for treatment effects in abstinence duration, analyses were conducted using Type III Wald chi-square tests in a generalized linear mixed (GLM) model with a negative binomial distribution, appropriate for non-normally distributed count variables and missing data across sessions. The model included treatment group and time as fixed effects, a random intercept for participant (to account for within-subject correlations across repeated measurements), and days between sessions (MRI2 and follow-up) as a covariate. A sensitivity analysis included a fixed effect of time (in days between sessions) coded as a continuous variable. All correlations that were conducted with abstinence duration at follow-up were corrected for abstinence at MRI2 and days between the MRI2 and follow-up visits.

## Results

### Clinical Outcomes

#### Abstinence duration

Participants in the MORE group demonstrated larger increases in abstinence duration from pre-treatment (M = 184.46, SD = 289.55) to post-treatment (M = 265.46, SD = 311.63) and follow-up (M = 435.54, SD = 414.88) as compared to those in the PST group [pre-treatment (M = 213.57, SD = 206.60), post-treatment (M = 252.91, SD = 238.42) and follow-up (M = 274.68, SD = 227.53)]. Corrected for days between sessions, the treatment group by session interaction was significant (χ² = 6.7, p = .035, Cohen’s w = .19), reflecting greater increases in abstinence in the MORE group relative to PST, driven by abstinence at follow-up. When the categorical session factor (MRI1, MRI2, follow-up) was replaced with days between sessions/visits as a continuous predictor, the model again revealed a significant treatment-by-time interaction (χ² = 5.28, p = .021, Cohen’s w = .17), confirming that abstinence duration increased more substantially in the MORE group compared with PST (Figure 2).

**Figure 2:**
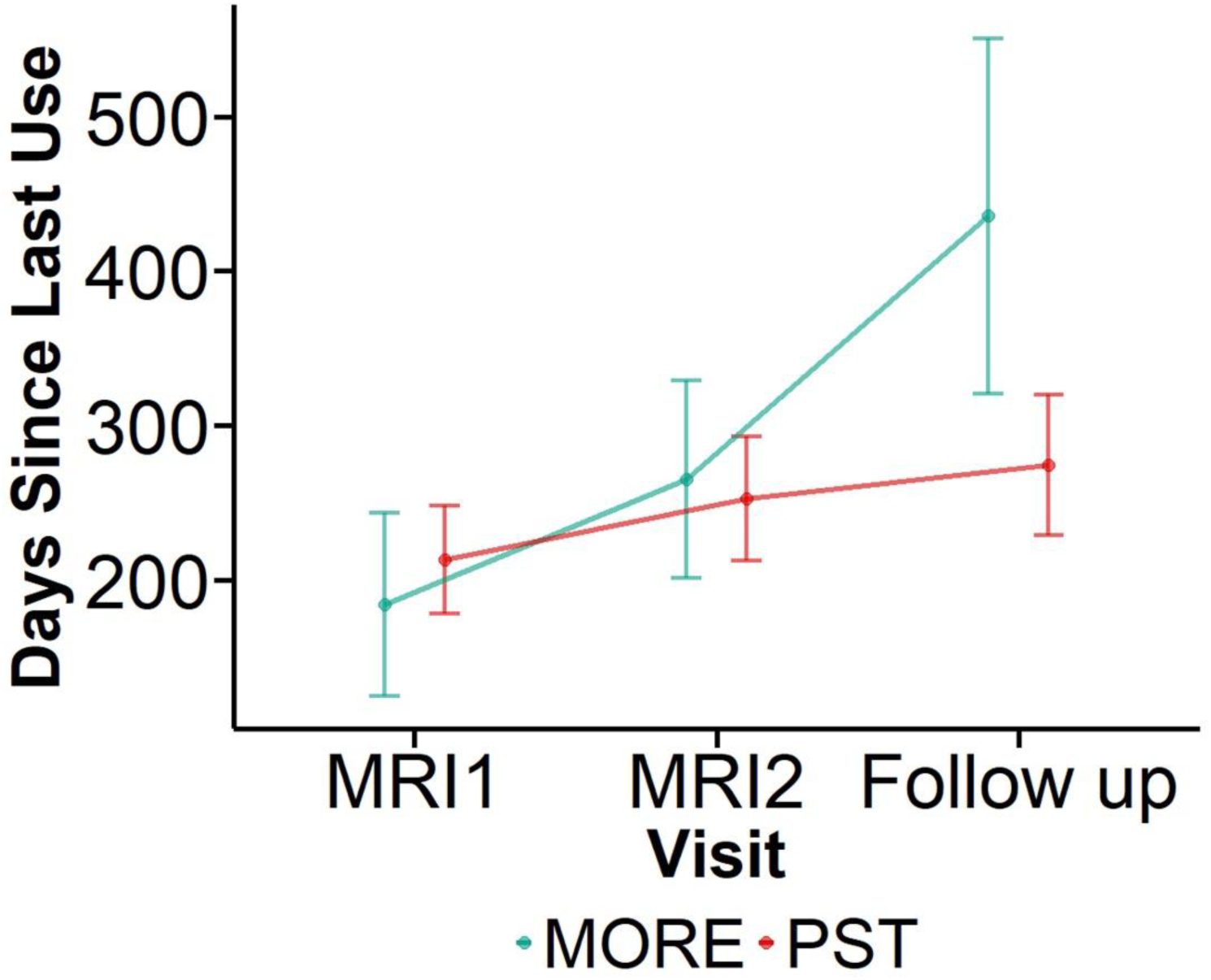
Abstinence Length between Treatment Groups Over Time. The numbers are represented in mean and standard errors. PST: Psychoeducational supportive therapy, MORE: Mindfulness-Oriented Recovery Enhancement; MRI1=pre-treatment scan; MRI2=post-treatment scan; Follow-up = approximately 4-months after MRI2.

#### Drug cravings

There were main effects of time for both cue-induced [F(1,56) = 11.63, p = 0.001, generalized eta-squared (ges) = 0.046] and scene-induced [F(1,34) = 10.13, p = 0.003, gest = 0.027] drug cravings with significant reductions of both craving measures after treatment across both groups. No significant main effects of treatment group or interaction effects were observed for both craving measures (p>0.115).

Planned within group paired t-tests showed the expected significant reductions in cue-induced cravings in both groups [MORE: t(23) = 2.32, p = 0.029, Cohen’s d = 0.48; PST: t(33) = 2.43, p = 0.021, Cohen’s d = 0.42]; yet scene-induced craving was reduced only in the MORE group [t(17) = 3.37, p = 0.0036, Cohen’s d = 0.79; PST: t(17) = 1.26, p = 0.223, Cohen’s d = 0.3].

We also examined whether treatment-related changes in these two craving measures were associated with abstinence length at follow-up within and across both groups. Using a significance threshold of p < 0.05/2=0.025 to correct for multiple comparisons, no significant associations emerged (ps > .025).

### BOLD fMRI Results

#### Cue-reactivity task

Whole-brain analyses revealed significant treatment group by session interaction effects in the right dlPFC/frontal eye field (Figure 3) across three emotion regulation-related contrasts: drug reappraisal > look drug, drug reappraisal > food savoring, and (drug reappraisal > look drug) > (food savoring > look food). These interaction effects were driven by the MORE group showing reductions in lateral PFC activity during drug reappraisal, compared to both passive drug cue viewing and food savoring, from pre-to post-treatment, whereas the PST group showed the opposite pattern (Figure 3A). In addition to the lateral PFC, regions within the default mode network encompassing the precuneus, posterior cingulate cortex, retrosplenial cortex, the lateral occipital cortex and pre/postcentral gyrus, showed similar treatment-related decreases in activity in the MORE group relative to the PST group (Table 2). See supplementary figure S3 for condition vs. implicit baseline breakdown.

**Figure 3:**
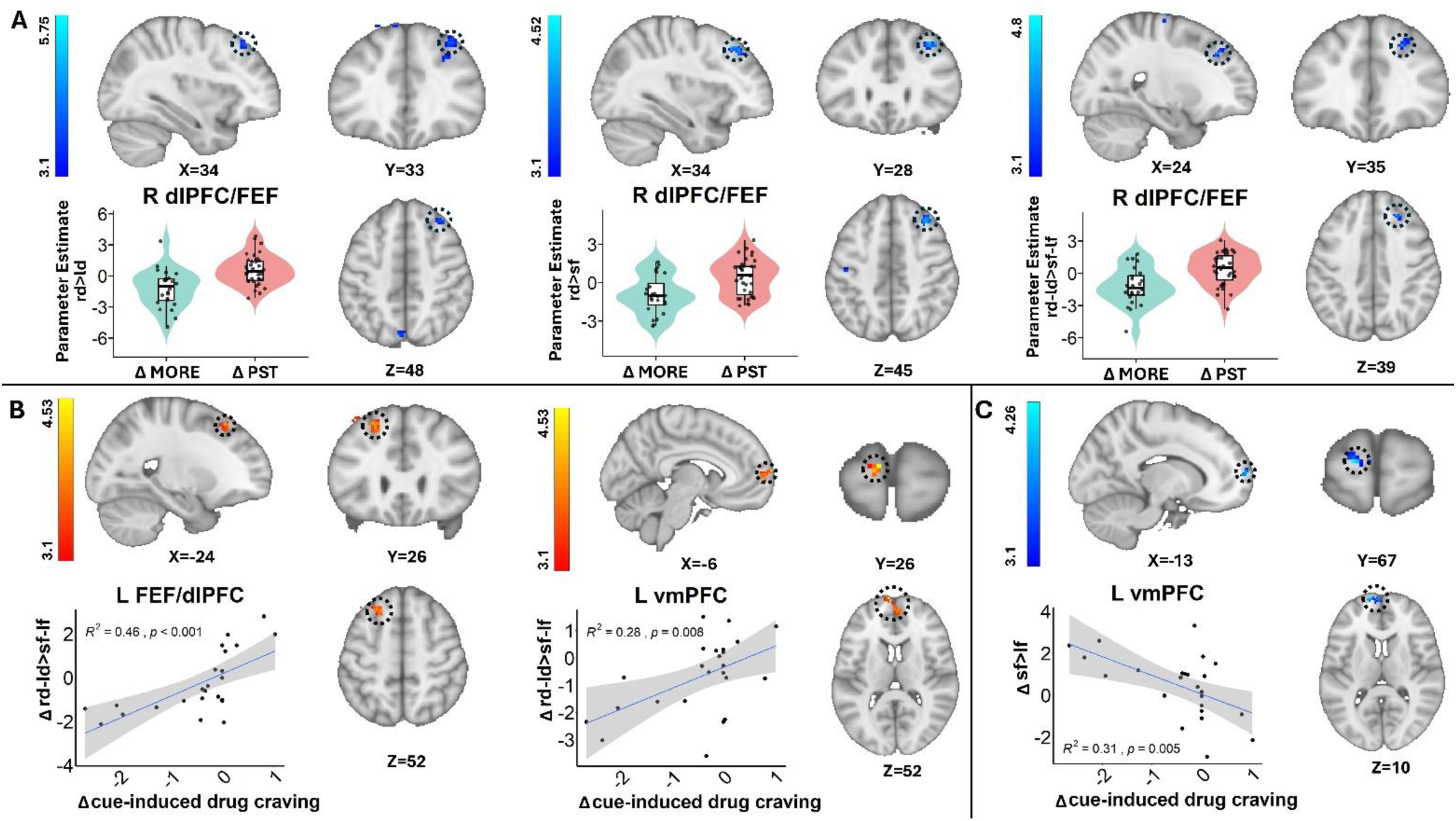
Treatment Effects on Prefrontal Cortex Activity during Emotion Regulation and their Correlations with Changes in Drug Craving. **A**: After treatment, the MORE group showed reduced activity in the dorsolateral prefrontal cortex (dlPFC)/frontal eye field (FEF) during drug cue reappraisal compared with drug cue passive viewing and food cue savoring (even when controlling for passive viewing), whereas the PST group showed the opposite pattern. **B**: In the MORE group, reductions in dlPFC and ventromedial prefrontal cortex (vmPFC) activity during drug cue reappraisal (relative to food cue savoring, controlling for passive viewing) were correlated with decreases in drug craving. **C**: In the MORE group, increases in vmPFC activity during food cue savoring were also correlated with reductions in drug craving. MORE: Mindfulness-Oriented Recovery Enhancement; PST: Psychoeducational supportive therapy; rd>ld=reappraise drug>look drug; rd>sf=reappraise drug>savor food, rd-ld>sf-lf=reappraise drug>savor food accounting for look drug and look food, L=left; R=right, Δ: differences between post- and pre-treatment measures. Cue-induced craving was measured from a post-MRI picture rating procedure. For visualization purposes, parameter estimates, depicting blood-oxygen-level-dependent signal, were extracted from corresponding FSL zstat images via 3-mm radius masks centered on Montreal Neurological Institute 152 coordinates from peak activity (black circles represent the approximate peak coordinates). Violin width reflects data density; boxes indicate the interquartile range with the median line, and points show individual participants. R^2^ and p values were derived from coordinates indicating peak effects.

**Table 2.**
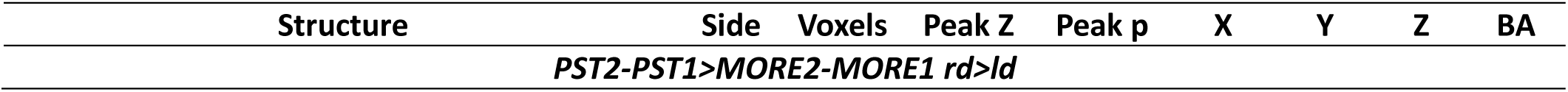
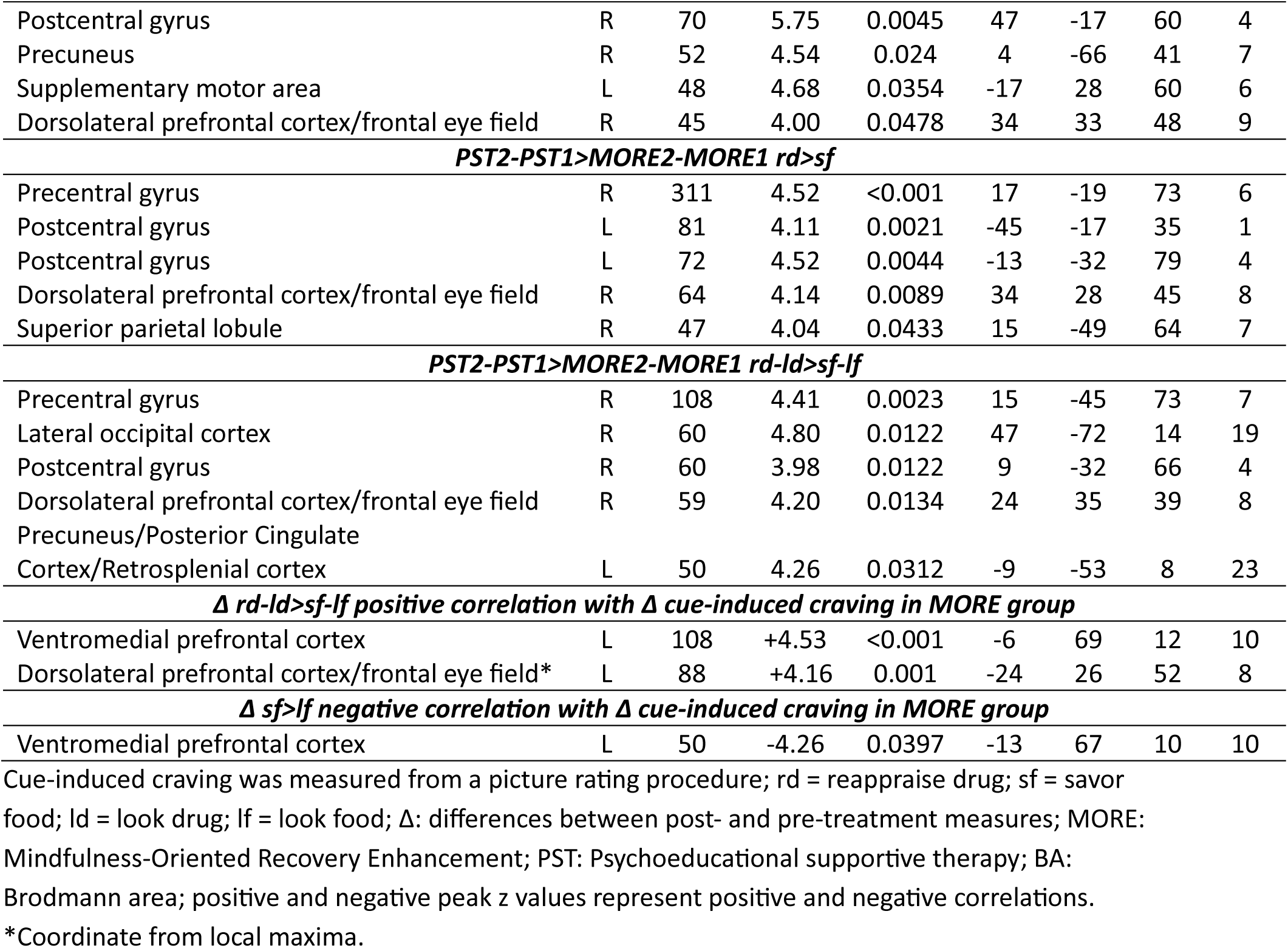
Coordinates for treatment effects and correlational results.

In whole-brain correlations between Δ brain activity and Δ drug craving, we observed significant positive correlations only in the MORE group, where reductions in dlPFC and vmPFC from pre-to-post-treatment during drug reappraisal (> food savoring, accounting for passive viewing of pictures matched on arousal) were associated with decreases in cue-induced drug craving (Figure 3B). These results suggest that the observed MORE-specific reduction in PFC activity during drug reappraisal vs. food savoring is meaningfully linked to lower cue-induced craving.

Furthermore, in the MORE group only, we found a significant negative correlation between savoring-related vmPFC activity and cue-induced drug craving, such that greater increases in vmPFC activation during food savoring (> look food) were associated with greater reductions in cue-induced drug craving following treatment with MORE (Figure 3C). This finding aligns with MORE’s therapeutic goal of reducing craving via shifting salience attribution away from drug-related cues and towards natural rewards by savoring their positive affective meaning.

We conducted two sets of correlation analyses examining abstinence length at follow-up: (1) whole-brain correlations with Δ brain activity within and across both groups, and (2) correlations using extracted signals from clusters that showed significant associations with Δ drug craving within the MORE group. Neither analysis yielded significant results.

#### Movie task

Compared with the PST group, the MORE group exhibited a significantly greater post-treatment reduction in drug cue-reactivity in an ROI on the border of the right ventromedial and dorsomedial PFC (vm/dmPFC) after FDR correction [MORE Δdrug bias = −1, PST Δdrug bias = 0.15, p=0.00019, q (FDR only over the PFC)=0.019, Figure 4].

**Figure 4:**
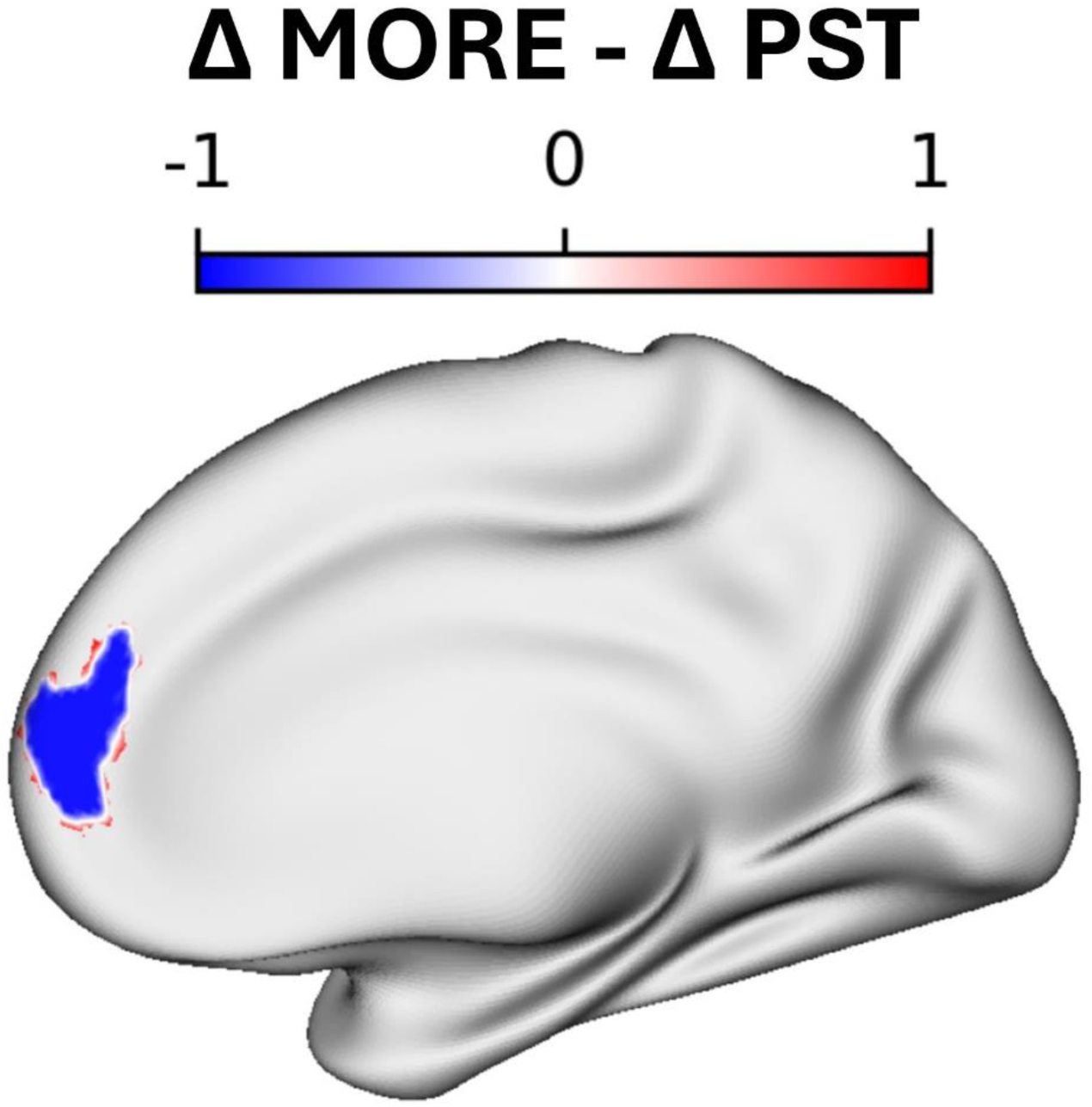
Treatment Effects on Naturalistic Drug-Biased Synchronization. After treatment the MORE group showed decreased movie drug cue-reactivity in a right vm/dmPFC ROI, while the PST group showed an increase. The color bar indicates the magnitude of difference in the Δ drug bias test statistic between groups. The drug bias statistic measures the fraction of TRs with a significant synchronized group response that followed drug content in the movie. Δ: post-minus pre-treatment drug bias measures; MORE: Mindfulness-Oriented Recovery Enhancement; PST: Psychoeducational supportive therapy

To assess whether responses in this vm/dmPFC region were related to drug craving, Δ ISC scores in this ROI were correlated with Δ scene-induced craving ratings. We found a significant positive correlation between Δ ISC and Δ scene-induced craving across all participants (spearman r=0.36, p=0.029), with no group differences in this correlation.

We also inspected the correlation between Δ ISC and abstinence length at follow-up within and across both groups, and did not find significant results (p > 0.13).

## Discussion

In this RCT, we demonstrated that, compared to PST, MORE produced significant improvements in both clinical and neural outcomes among individuals with OUD receiving inpatient treatment. Crucially, participants randomized to eight weeks of MORE achieved significantly greater increases in abstinence duration ∼4-months following treatment than those in PST, and, in within-group analyses, reported significant decreases in scene-induced drug craving. These results are consistent with prior evidence supporting MORE’s efficacy as a treatment for opioid misuse and OUD^42^, reducing rates of return to opioid use and treatment dropout^20^, whereby reductions in opioid craving and opioid misuse were attributed to increases in EEG responses and positive affective reactivity to alternative reward savoring.^22,43^ Importantly, we can now link these clinical therapeutic effects of MORE to specific neural changes and a putative mechanism whereby MORE recalibrates the neurobiological processes underlying the regulation of drug cue-reactivity, representing a novel advance in the treatment of OUD and, more broadly, substance use disorders. Specifically, participants in MORE exhibited reductions in activity in the dlPFC/frontal eye fields during reappraisal of drug-related cues compared to their passive viewing and savoring of food cues. Similar reductions were observed in posterior regions of the default brain network (e.g., the precuneus). Whole-brain voxel-wise analyses revealed that, within the MORE group, greater decreases in dlPFC and vmPFC activity during drug cue reappraisal (relative to food savoring), as well as greater increases in vmPFC activation during savoring of natural reward cues (relative to their passive viewing), were associated with larger reductions in cue-induced drug craving. Furthermore, when participants viewed a naturalistic drug-themed movie, compared to PST, those in the MORE group exhibited greater post-treatment reduction in drug-biased brain synchronization within the vm/dmPFC, suggesting diminished preferential engagement of limbic frontal networks by real-life dynamic drug cues. This post-treatment reduction of vm/dmPFC synchronization was associated with larger decreases in scene-induced drug craving (across both groups), again underscoring the clinical relevance of these neural changes. To our knowledge, this is the first neuroimaging RCT to assess the effects of a mindfulness-based intervention on the neural substrates underlying regulation of drug cue-reactivity in addiction.

Our finding that MORE reduced lateral PFC activity during drug reappraisal (using static images) aligns with and extends prior evidence that mindfulness-based interventions modulate neurophysiological reactivity to drug cues in people with substance use disorders. In people with alcohol use disorder, treatment with Mindfulness-Based Relapse Prevention was associated with decreased alcohol cue-reactivity as indexed by LPPs.^44^ Similarly, in a randomized EEG study, patients with opioid misuse who underwent eight weeks of MORE exhibited reduced LPP during reappraisal of opioid cues as compared to passive viewing of those cues.^21^ The present study advances this literature by identifying underlying neural substrates implicated in these earlier findings. Specifically, our results highlight the role of the dlPFC/frontal eye field, regions consistently implicated in the deployment of top-down attentional and regulatory control over salient stimuli.^45–47^ This interpretation is consistent with broader cognitive neuroscience evidence linking these regions to emotion regulation processes,^48,49^ with prior work using Bayesian inference showing their selective involvement during reappraisal.^50^

In our previous report with a similar iOUD sample and compared to healthy controls, we observed heightened lateral PFC and inferior frontal gyrus activity during drug cue reappraisal, interpreted to reflect inefficient or compensatory deployment of cognitive control resources.^12^ In particular, this dlPFC drug-biased reappraisal hyperactivity (relative to food savoring and passive drug cue viewing, respectively) was positively associated with methadone dose (a measure of addiction severity) and negatively with craving.^12^ Building on these previous findings, the present trial demonstrates that this neural dysfunction is malleable to treatment with MORE. That is, iOUD in the MORE group exhibited reduced reappraisal-related cognitive control network reactivity, and when compared to food savoring, this relative reduction in reappraisal-related activation was linked with treatment-associated reductions in cue-induced craving. Similar decreases were noted in the default mode network encompassing the precuneus, suggesting lower drug cue induced rumination ^51^. The vmPFC, another core region in this network, similarly showed normalization of function whereby in contrast to our prior report (of drug cue hyperactivations in this region as associated with increased craving)^12^, in this study its reduced drug reappraisal and enhanced food savoring activity correlated with reduced craving with treatment. The present results are consistent with an earlier pilot quasi-experimental fMRI study in smokers, which showed that MORE decreased medial PFC and ventral striatal reactivity to smoking cues, while enhancing mPFC-striatal responses to natural reward cues during savoring.^52^ Taken together, the observed dlPFC and vmPFC effects indicate that MORE normalizes hyper-reactivity to drug cues and hypo-reactivity to natural rewards, facilitating a shift from drug-biased to non-drug salience processing, in line with the iRISA model of addiction^10^ and MORE’s hypothesized role in restructuring reward processes.^53^

In the movie paradigm, MORE elicited a significant reduction in drug-biased synchronization within the limbic vm/dmPFC. Inter-subject synchronization quantifies shared neural dynamics across individuals exposed to the same stimuli.^54^ The observed decrease in vmPFC synchronization following MORE may signify diminished collective craving-related neural engagement, and indeed reductions in the overall ISC within this region were associated with decreases in movie scene-induced drug craving. Given vmPFC’s established role in value assignment^55^, and prior evidence that synchronized vmPFC activity tracks affective valuation during naturalistic viewing^56^, these findings further suggest that MORE supports the regulatory resources that rebalance drug-biased salience attribution in people with drug addiction. Taken together, and while lending enhanced validity and reliability to our findings, our dual static (picture viewing) and narrative-based (i.e., movie watching) approaches to estimating changes in drug cue-reactivity may also inform distinct MORE-induced recovery mechanisms. Indeed, the cue-reactivity task with static images explicitly instructed participants to downregulate responses to drug cues and upregulate responses to food cues, whereas no such instructions were provided during movie watching. Explicit (i.e., instructed) and implicit (i.e., spontaneous) emotion regulation have been preferentially associated with lateral and medial PFC engagement, respectively.^57^ Potentially, as MORE reduces the incentive salience of drug cues and cravings,^18,58,59^ the need to engage neural regions associated with both explicit and implicit emotion regulation strategies may be attenuated. Interestingly, using the cue-reactivity task we did not observe group-level changes in conventional passive cue-reactivity contrasts (e.g., look drug vs. look food/neutral). This null result may reflect more specificity of the therapeutic effects of MORE on the proactive regulation (and not mere passive processing) of drug cue-reactivity or reflect the mixed emotion regulation conditions used to down-regulate drug cue-reactivity in this picture task; these results may also suggest that naturalistic drug cues (movie scenes) are more sensitive for detecting early recovery of bottom-up/limbic drug-biased salience attribution. Therefore, our design may capture unique components of the recovery of salience attribution in drug addiction, offering mechanistic insight into how mindfulness training reshapes (both top-down and bottom-up) neural responses to drug vs. non-drug rewards.

Several study limitations and future directions should be noted. First, larger samples are needed to inspect individual differences in MORE’s therapeutic effects; for example, we did not observe significant correlations with abstinence length at follow-up potentially due to participants dropout (MORE: n=24 to 13; PST: n=35 to 25). Additionally, we did not examine sex differences despite recent evidence that drug cue-reactivity is instantiated differently in men and women with addiction^32^. Because in our prior study we also found effects of ovarian hormones (i.e., estradiol and progesterone) in the lateral and medial PFC during drug cue-reactivity and its regulation via reappraisal, the same regions modulated by MORE, future efforts could explore the potential optimization of treatment effects by their timely (menstrual phase-locked) delivery. Second, our sample only comprised inpatient iOUD receiving medications for OUD. Given morphometric differences reported between opioid (e.g., heroin) and stimulant (e.g., cocaine) addiction, particularly within prefrontal circuitry^60^, future work should extend these findings across substance classes and medication status, to outpatient settings, and across psychosocial treatments, to determine substance-specific and treatment effects. Third, dose-response relationships could not be evaluated here; future studies should test treatment duration and practice dose (e.g., MORE session frequency) as predictors of neural and clinical change. Finally, future work should incorporate functional connectivity analyses, which may help explain the absence of ventral striatal effects in the current study. For example, although the nucleus accumbens showed cue-reactivity in our prior work,^12^ its involvement here may be obscured at the activation level and could be revealed by analyses emphasizing prefrontal-striatal connectivity.

In summary, this RCT provides the first evidence that MORE modulates prefrontal mechanisms of drug cue-reactivity, salience attribution, and cognitive control in opioid addiction. MORE reduced within-group drug craving and extended abstinence length, underscoring its clinical utility in combating the ongoing opioid crisis. Crucially, the current study shows that MORE attenuated prefrontal activation during drug cue regulation, decreasing drug-biased prefrontal group-level/shared synchronization in naturalistic contexts, with reductions in drug bias during reappraisal and enhancements in alternative reward savoring linked to reductions in craving. Together, these findings suggest that MORE may recalibrate neural systems governing reward valuation and self-regulation, consistent with the iRISA model, and highlight MORE’s promise as a biologically grounded adjunct to pharmacotherapy for OUD. Future efforts could refine these neural targets (e.g., via neuromodulatory strategies including non-invasive brain stimulation, neurofeedback, or just-in-time digital delivery of MORE) to accelerate the translation of mindfulness-based treatments into precision addiction medicine.

## Author Contributions

Study design: AOC, ELG, RZG.

Data acquisition: YH, AOC, GK, SGK, NEM, KRD, YYW, MB.

Data preparation, analysis, and visualization: YH, GK, KRD.

Initial manuscript writing: YH, AOC.

Manuscript review and revision: All authors.

Treatment training and delivery: ERB, POG, NAK, ELG. Funding acquisition: RZG.

## Data Availability

All data produced in the present study are available upon reasonable request to the authors

