## supplementary material for "Mindfulness-Oriented Recovery Enhancement rebalances prefrontal responses to drug and natural reward cues in opioid use disorder"

#### Exclusion criteria

Exclusion criteria for participants were the following: 1) DSM-5 diagnosis for schizophrenia or developmental disorder; 2) Head trauma with loss of consciousness (>30 min); 3) History of neurological disease of central origin; 4) Cardiovascular, metabolic, endocrinological, oncological, autoimmune, and active infectious diseases including Hepatitis B and C or HIV/AIDS; 5) Metal implants or other magnetic resonance imaging (MRI) contraindications (including pregnancy); 6) Court mandated treatment. We did not exclude for DSM-5 diagnosis of a drug use disorder other than opiates as long as heroin was the primary drug of choice/reason for treatment-seeking since individuals with heroin use disorder commonly use alcohol, amphetamines, benzodiazepines, other sedatives, cocaine, and marijuana in addition to heroin.

### Participants

All 59 participants who were involved in the main fMRI analyses met criteria for OUD. Other comorbidities included major depressive disorder (n=27), posttraumatic stress disorder (n=12), panic disorder (n=8), general anxiety (n=5), agoraphobia (n=2), obsessive compulsive disorder (n=2), eating disorder (n=1), cocaine use disorder (n=19), other than crack/cocaine stimulants use disorder (n=2), alcohol use disorder (n=14), tranquilizer use disorder (n=10), and cannabis use disorder (n=3).

Primary routes of opioid administration were intravenous injection (n=28), intranasal (n=27), smoking/inhaling (n=2), non-intravenous injection (n=1), and oral (n=1).

Across all the participants, urine toxicology assessment on the first MRI visit indicated the presence of fentanyl (n=2), marijuana/THC (n=1), cocaine (n=2), methamphetamine (n=1), methadone (n=52), buprenorphine (n=6), opioids (n=2), and antidepressants (n=26). Eight individuals reported opioid use during the 30 days preceding the scan.

At the second MRI, urine toxicology assessment indicated the presence of fentanyl (n=4), marijuana/THC (n=1), cocaine (n=2), morphine (n=3), benzodiazepines (n=1), methadone (n=51), buprenorphine (n=6), opioids (n=2), and antidepressants (n=29). One datapoint for urine toxicology data was missing. Eight individuals reported opioid use during the 30 days preceding the scan.

Of the 59 participants included in the main analysis, 40 returned for the follow-up visit and 38 provided analyzable data. In these participants, urine toxicology assessment indicated the presence of fentanyl (n=2), cocaine (n=2), amphetamine (n=1), methamphetamine (n=1), morphine (n=2), benzodiazepine (n=1), methadone (n=29), buprenorphine (n=3), opioids (n=3), and antidepressants (n=20). Five individuals reported opioid use during the 30 days preceding the follow-up visit.

### MRI data acquisition

The MRI protocol was optimized to be Human Connectome Project compatible[1] and the data was collected on a Siemens 3-T Skyra scanner (Siemens Healthcare, Erlangen, Germany) using a 32-channel head coil. Anatomical T1-weighted structural images were acquired using the following parameters: 3D MPRAGE (Magnetization-Prepared Rapid Gradient-Echo) sequence with FOV of 256 × 256 × 179 $mm^3$, 0.8 mm isotropic resolution, TR/TE/TI = 2400/2.07/1000 ms, 8° flip angle with binomial (1, −1) fat saturation, 240 Hz/pixel bandwidth, 7.6 ms echo spacing, and in-plane acceleration (GRAPPA-generalized autocalibrating partially parallel acquisitions) factor of 2, with a total acquisition time of approximate time of 7 minutes. The blood-oxygen-level-dependent (BOLD) fMRI responses were measured as a function of time using T2*-weighted single-shot multiband accelerated (factor of 7) gradient-echo echo-planar image (EPI) sequence [TE/TR=35/1000 ms, 2.1 isotropic mm resolution, 70 axial slices without gaps for the whole brain (147mm) coverage, FOV 206 × 181 mm, matrix size 96 × 84, 60°-flip angle (approximately Ernst angle), blipped CAIPIRINHA (Controlled Aliasing in Parallel Imaging Results in Higher

Acceleration) phase-encoding shift=FOV/3, 1860 kHz/Pixel bandwidth with ramp sampling, echo spacing 0.68 ms, and echo train length 84 ms]. In addition to the picture cue-reactivity task and movie watching task, the 2-hour scan included additional structural and functional procedures reported elsewhere.[2–8]

### Picture Cue-reactivity task

#### Task instructions

Task instructions for the look, reappraise, and savor conditions were adapted from previous studies[9,10]. During the look condition, participants were instructed to “keep viewing the picture normally”. During the reappraise condition, participants were instructed to reduce their emotional reactivity to the drug pictures in three practice trials, each providing a different strategy: 1) “Try to imagine that the scenario is not real, that it is from a movie, and these are all actors”; 2) “Try to imagine that the heroin is not real, that it is just a prop”; 3) “You can focus on how this is just a picture, and tell yourself that it is not real heroin”. During the savor condition, participants were instructed to increase their emotional reactivity to the food pictures in three other practice trials, each providing a different savoring strategy: 1) “You can imagine that you are holding the food in the picture, and feeling the weight of it in your hands, enjoying its pleasant smell”; 2) “You can focus on how good the food looks or imagine how good it would taste, savoring the delicious taste of the food”; 3) “Imagine the sensation of how it would feel in your mouth or how it feels once you’ve eaten it”. Participants were instructed to verbally describe their reappraisal and savoring strategies during these practice trials to ensure task comprehension, but they were instructed to refrain from speaking during the fMRI task trials in the scanner.

#### Pre- and post-cue-reactivity task and post-MRI ratings

Participants provided drug and food craving ratings, as well as a ratings of motivation to complete the task on a 10-point scale immediately before the functional MRI (fMRI) cue-reactivity task (i.e., “Please rate how strong your desire for heroin is currently on a scale of 0-9”, “Please rate how strong your desire for food is currently on a scale of 0-9”, and “Please rate your motivation to complete this task on a scale of 0-9”) (i.e., baseline). Immediately after the cue-reactivity task, in addition to the same three questions, participants were asked to provide self-evaluation of the difficulty and effectiveness of their reappraisal and savoring performance on a 10-point scale (i.e., “How difficult did you find it to decrease your emotional reactivity to the heroin pictures during this task?”, “How well do you think you decreased your emotional reactivity to the heroin pictures during this task?”, “How difficult did you find it to increase your emotional reactivity to the food pictures during this task?” and “How well do you think you increased your emotional reactivity to the food pictures during this task?”).

Post-MRI, we also acquired ratings for half of the images viewed during the fMRI task. These images were pseudorandomized by subject and session. For valence ratings, participants were asked ‘How pleasant do you find the above picture?' on a 5-point scale from 1 (very unpleasant) to 5 (very pleasant). For arousal, participants were asked "How emotional do you feel about the above picture?" on a 5-point scale from 1 (calm, no emotion) to 5 (extremely emotional). For wanting, participants were asked "How strong is your desire to use the above substance (or food)?" on a 5-point scale. The latter drug rating was used as our a priori selected cue-induced drug craving rating (see main text).

#### Treatment effects

To analyze pre- and post-task drug/food craving ratings between treatment groups before and after treatment, a 2 (treatment group: MORE/PST) by 2 (session: MRI1/MRI2) by 2 (task: pre-/post-task) by 2 (cue: drug/food) mixed analysis of variance (ANOVA) was conducted. We found significant main effects of task [post>pre: $F(1,52) = 8.61$, $p = 0.005$, generalized eta-squared (ges) = 0.011], cue [food>drug: $F(1,52) = 79.82$, $p < 0.001$, ges = 0.286] and session [MRI1>MRI2: $F(1,52) = 16.99$, $p < 0.001$, ges = 0.040] and interaction effect between treatment group and cue [MORE>PST in overall drug craving: $F(1,52) = 11.48$, $p = 0.001$, ges = 0.054]. No other significant effects were found (ps > 0.127).

Because these craving ratings did not show a treatment group × session interaction, the between-group difference in overall drug craving is unlikely to account for the observed treatment group × session interactions in the right dlPFC/FEF reappraisal activity. Consistent with this interpretation, whole-brain voxel-wise correlations (with $Z > 3.1$ and $p < 0.05$) did not show significant results for the pre-treatment pre-task (baseline) drug craving (with treatment-related changes in the cluster of right dlPFC/FEF reappraisal activity) in either group.

A 2 (treatment group: MORE/PST) by 2 (session: MRI1/MRI2) by 2 (task: pre/post) mixed ANOVA for motivation ratings showed a significant main effect of treatment group [PST>MORE: $F(1,52) = 6.20$, $p = 0.016$. ges = 0.058] and interaction effect between treatment group, task, and session [MRI2>MRI1 in post in MORE and PST>MORE in post in MRI1 and in pre in MRI2, $F(1,52) = 5.02$, $p = 0.029$, ges = 0.012]. No other significant effect was observed (ps > 0.345).

Regarding self-evaluated emotion regulation performance, four 2 (treatment group: MORE/PST) by 2 (session: MRI1/MRI2) mixed ANOVAs for difficulty and effectiveness were conducted for drug reappraisal and food savoring separately. We found significant main effect of treatment group for drug reappraisal difficulty [MORE>PST: $F(1,53) = 5.05$, $p = 0.029$, ges = 0.061], drug reappraisal effectiveness [PST>MORE: $F(1,53) = 5.38$, $p = 0.024$, ges = 0.042], and food savoring difficulty [MORE>PST: $F(1,53) = 4.76$, $p = 0.034$, ges = 0.042]. No other significant effect was observed (ps > 0.280).

Correlations between pre-treatment motivation (baseline), drug reappraisal difficulty and effectiveness, and food savoring difficulty with MRI2-MRI1 changes (deltas) in the extracted right dlPFC/FEF ROI showing reappraisal activity across all three contrasts were not significant (ps > 0.157).

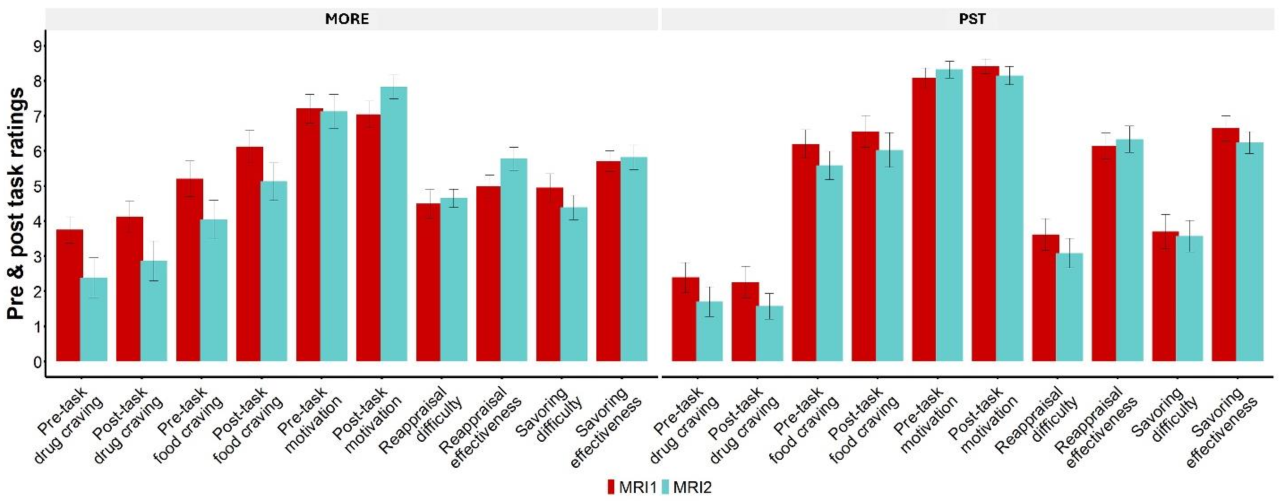


**Figure S1**. Pre- and post-cue-reactivity task ratings.

To test potential treatment effects between the valence and arousal ratings for food, drug and neutral images, two separate 2 (treatment group: MORE/PST) by 2 (session: MRI1/MRI2) by 3 (images: drug/food/neutral) mixed ANOVAs were conducted. For valence, there were significant main effects of image [food>neutral>drug: $F(1.29, 72.2) = 145.76$, $p < 0.001$, ges = 0.546] and session (MRI1>MRI2: $F(1,56) = 10.42$, $p = 0.002$, ges = 0.023]. No other significant effects were observed ($p > 0.097$). Similarly, for arousal, there were significant main effects of image [food=drug>neutral: $F(1.2, 67.03) = 13.00$, $p < 0.001$, ges = 0.092] and session (MRI1>MRI2: $F(1,56) = 6.00$, $p = 0.017$, ges = 0.014]. No other significant effects were observed ($p > 0.129$).

A 2 (treatment group: MORE/PST) by 2 (session: MRI1/MRI2) by 2 (image: drug/food) mixed ANOVA on the wanting/cue-induced craving ratings again showed similar significant main effects for image [food >drug: $F(1,56) = 41.38$, $p < 0.001$, ges = 0.180] and session (MRI1>MRI2: $F(1,56) = 10.88$, $p = 0.002$, ges = 0.030]. No baseline differences in cue-induced drug craving ($p = 0.15$) and no other significant effects were observed ($p > 0.255$).

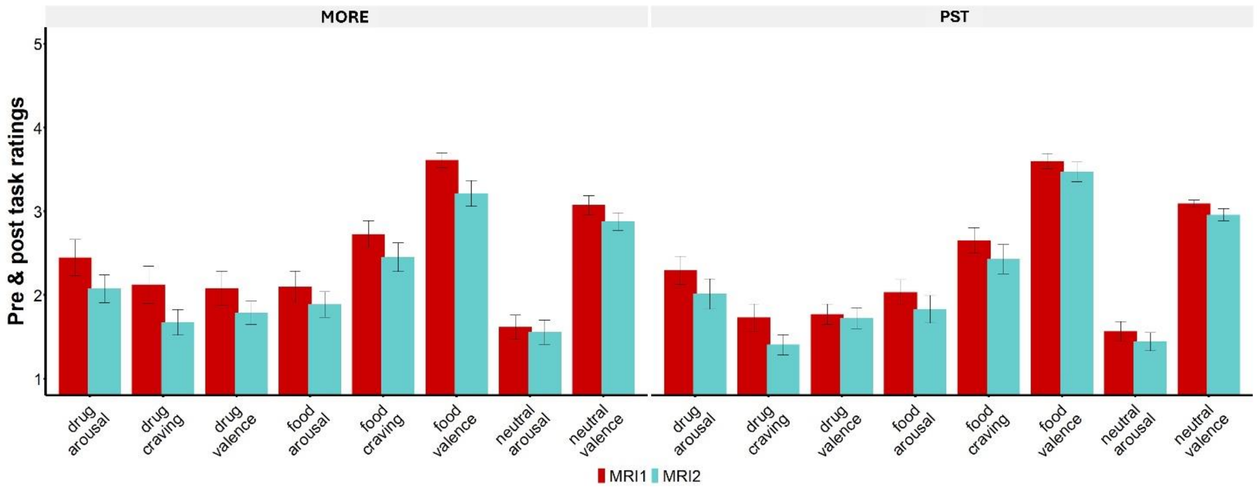


**Figure S2**. Post-cue-reactivity task picture ratings.

### MRI preprocessing

Raw BOLD-fMRI data in DICOM (Digital Imaging and Communications in Medicine) format were first converted to NIFTI (Neuroimaging Informatics Technology Initiative) via dcm2nii[11] and HeuDiConv (https://github.com/nipy/heudiconv) and adapted to the BIDS (Brain Imaging Data Structure) format[12] and then preprocessed via fMRIprep pipeline (version 20.2.1).[13,14] The structural images were Intensity-normalized and skull-stripped with ANTs (Advanced Normalization Tools).[15,16] Volume-based spatial normalization through nonlinear registration into ICBM (International Consortium for Brain Mapping) 152 Nonlinear Asymmetrical template was performed with ANTs.[15,17] Brain tissue was segmented into the cerebrospinal fluid, white matter, and gray matter through FSL's [Functional MRI of the Brain (FMRIB) Software Library] FAST (FMRIB Automated Segmentation Tool).[18] Susceptibility distortion correction using echo-planar field maps acquired in opposing phase-encoding directions was applied to the functional images via 3dQwrap in AFNI (Analysis of Functional Neuro Images).[19] Motion artifacts were estimated and corrected for the functional images via FSL's MCFLIRT (FMRIB Linear Image Registration Tool with motion correction).[20] Motion- and distortion-corrected images were then co-registered to participants' structural T1w images with the boundary-based registration with 9 degrees of freedom using FSL'S FLIRT,[17,21] and normalized to ICBM 152 nonlinear asymmetrical template.[17] Individual anatomical T1 images were underwent Freesurfer's recon-all,[22] prior incorporated into fMRIprep's preprocessing pipeline to improve registration for the functional image. One MRI2 scan from the PST group did not include this step due to poor surface parcellation outcome. All preprocessed functional images were visually inspected.

In addition to fMRIprep, we identified volumes with spikes in translation and rotation parameters using a typical boxplot threshold (75th percentile + 1.5 times the interquartile range) in relation to a reference image volume using FSL's fsl_motion_outlier. On average, we regressed out 5.7% of the total volumes in each run (range: 1.32%-12.5%) for the baseline fMRI and 5.9% of the total volumes in each run (range:

1.32%-12.7%) for the post-treatment fMRI. There were no significant differences in the percentage of motion outliers between MORE (mean = 5.8 ± 2.1) and PST (mean = 5.6 ± 2.4; W = 470.5, p = 0.44) in the baseline fMRI. Additionally, the frame-wise displacement showed no significant differences between MORE (mean 0.283 ± 0.118) and PST (mean = 0.314 ± 0.148; W = 466, p = 0.49) in the baseline fMRI. For the post-treatment fMRI, there were no significant differences in the percentage of motion outliers between the MORE (mean = 5.5 ± 2.2) and PST MRI (mean = 6.2 ± 2.1; $t_{57}$ = 1.12, p = 0.27). Similarly, the frame-wise displacement did not significantly differ between the MORE (mean = 0.301 ± 0.100) and PST (mean = 0.345 ± 0.146; W = 465, p = 0.50) at the post-treatment fMRI. A high-pass filter (100 s cutoff) was applied to the functional data to ignore scanner drift. Lastly, the preprocessed data were spatially smoothed with a Gaussian kernel (5-mm full-width at half maximum) to improve signal-to-noise ratio.

### Movie task

#### Movie ratings and scene-specific post-movie survey

Immediately before and after the movie, subjects were asked to rate their desire for heroin on a scale of 0-9. There were significant main effects of session (MRI1>MRI2: F(1, 32) = 11.05.76, p = 0.002, ges = 0.056) and task (post-task>pre-task: F(1,32) = 9.09, p = 0.005, ges = 0.014). No other significant effect was found (ps > 0.08). Within 45 min after movie viewing, participants also completed a scene-specific craving survey following our previously published procedure.[23] Briefly, 34 three-second clips were extracted at 30 s intervals from the 17-min movie. For each clip, participants rated their craving as experienced during in-scanner viewing. The survey also included comprehension, attention, and memory questions related to the movie watching experience. There were no group differences in these measures or their deltas between sessions (ps > 0.18).

#### Anatomical data preprocessing with fMRIprep version 20.2.1[13,14]

For each subject, a T1-weighted (T1w) image was corrected for intensity non-uniformity with N4BiasFieldCorrection[24], distributed with ANTs 2.3.3[25] and used as T1w-reference throughout the workflow. The T1w-reference was then skull-stripped with a Nipype implementation of the antsBrainExtraction.sh workflow (from ANTs), using OASIS30ANTs as the target template. Brain tissue segmentation of cerebrospinal fluid, white-matter and gray-matter was performed on the brain-extracted T1w using FAST [FSL 5.0.9, RRID:SCR_002823[26]]. Volume-based spatial normalization to a standard space (MNI152NLin2009cAsym) was performed through nonlinear registration with antsRegistration (ANTs 2.3.3), using brain-extracted versions of both T1w reference and the T1w template. The ICBM 152 Nonlinear Asymmetrical template version 2009c[27] [RRID:SCR_008796; TemplateFlow ID: MNI152NLin2009cAsym] was used for spatial normalization.

#### Functional data preprocessing with fMRIPrep version 20.2.1

For all BOLD movie runs (across all subjects), the following preprocessing was performed. First, a reference volume and its skull-stripped version were generated by aligning and averaging a single-band reference. A B0-nonuniformity map (or fieldmap) was estimated based on two echo-planar imaging (EPI) references with opposing phase-encoding directions, with AFNI's 3dQwarp[28]. Based on the estimated susceptibility distortion, a corrected EPI (echo-planar imaging) reference was calculated for a more accurate co-registration with the anatomical reference. The BOLD reference was then co-registered to the T1w reference using FLIRT [FSL 5.0.9[29]] with the boundary-based registration[30] cost-function. Co-registration was configured with nine degrees of freedom to account for distortions remaining in the BOLD reference. Head-motion parameters with respect to the BOLD reference (transformation matrices, and six corresponding rotation and translation parameters) were estimated before any spatiotemporal filtering using MCFLIRT [FSL 5.0.9[31]]. First, a reference volume and its skull-stripped version were generated using a custom methodology of fMRIPrep. The BOLD time-series were resampled onto their original, native space by applying a single, composite transform to correct for head-motion and susceptibility distortions. The BOLD time-series were resampled into the MNI152NLin2009cAsym standard space and used for further custom preprocessing.

#### Custom preprocessing

We applied the same preprocessing pipeline as in our prior work using this task.[23] In summary, spatially normalized BOLD data (via MNI152NLin2009cAsym) were masked to gray matter, binarized with thresholder at 95% probability, smoothed (via 6 mm FWHM), and had the first 10 samples removed. [32] Motion parameters (translations/rotations, their squares, derivatives, and squared derivatives) and the cerebrospinal fluid component from fMRIPrep were regressed out. The high-pass filtering (140 s), linear detrending, and z-scoring were further regressed out using nilearn.signal.clean.[33]

#### Global component and selective components

For each subject, the preprocessed BOLD time series were averaged over all gray-matter voxels and then z-scored, resulting in a single global component per subject. Selective components were then derived by regressing the time series at each voxel within a subject onto the global component for that subject with a least-squares linear model. The residual of this linear fit was then z-scored and kept as the selective component for that subject-voxel.[34]

#### Parcellations and shared response model

Cortical ROIs were selected by applying the Schaefer functional parcellation[35] with 400 regions. Subcortical ROIs were selected with the Melbourne Subcortical Atlas[36] with 50 regions. To simplify analyses and focus on shared group responses, we derived a single time series per ROI for each subject by applying a shared response model with a single shared component per group[37]. This method projects the data for each subject to a single component that maximally explains the observed voxel-wise data within each ROI via a linear transform. The shared response model was implemented via the BrainIAK toolbox[38]. All further analyses were conducted on this shared component within each ROI.

### Supplementary Figure

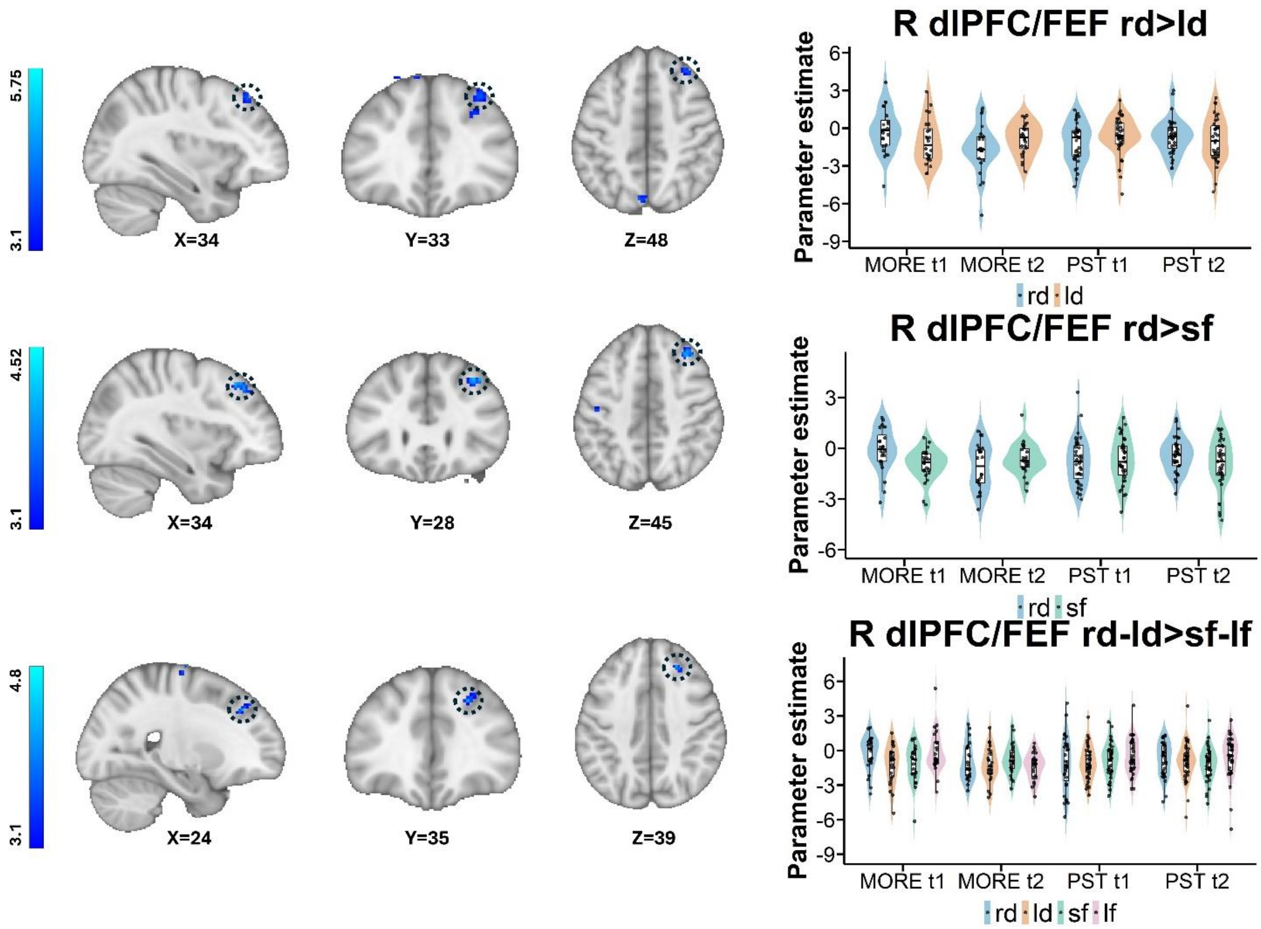


**Figure S3: Treatment Effects on Prefrontal Cortex Activity during Emotion Regulation vs. Implicit Baseline**

MORE: Mindfulness-Oriented Recovery Enhancement; PST: Psychoeducational supportive therapy; t1=; t2=post-treatment scan; dlPFC=dorsolateral prefrontal cortex; FEF=frontal eye field; rd>ld=reappraise drug>look drug; rd>sf=reappraise drug>savor food, rd-ld>sf-lf=reappraise drug>savor food accounting for look drug and look food, R=right For visualization purposes, parameter estimates, depicting blood-oxygen-level-dependent signal, were extracted from corresponding FSL zstat images via 3-mm radius masks centered on Montreal Neurological Institute 152 coordinates from peak activity (black circles represent the approximate peak coordinates). Violin width reflects data density; boxes indicate the interquartile range with the median line, and points show individual participants.
